# Cross-database adaptation of machine learning models for predicting sustained antibiotic exposure among patients with intracerebral hemorrhage in intensive care units

**DOI:** 10.64898/2026.10.11.26365235

**Authors:** Sakshie Pathak, Yong Si, Janet Sanjaya, Mohammadsaeed Haghi, Nausin Tabassum Kudrot, Greg Placencia, Kamiar Alaei, Maryam Pishgar

## Abstract

**Background:** Patients with intracerebral hemorrhage (ICH) in intensive care units (ICUs) frequently receive antibiotics early in critical illness. Such exposure reflects suspected infection, physiological deterioration, monitoring intensity, and clinical management and may vary across institutions and electronic health record databases. We evaluated target-domain adaptation for modeling sustained antibiotic exposure in critically ill patients with ICH.

**Methods:** We analyzed derived ICH cohorts from MIMIC-IV and the eICU Collaborative Research Database. MIMIC-IV included 2,024 ICU stays and served as the source cohort. The eICU cohort comprised 1,277 stays and was divided, at the ICU-stay level, into a development subset (766 stays) and a held-out evaluation subset (511 stays). The eICU outcome was sustained antibiotic exposure on at least two ICU days. Candidate models incorporated demographic, comorbidity, physiological, laboratory, neurologic, respiratory, temporal, and measurement-process features. Timestamped observations for static and temporal tabular families were censored at the recorded first-antibiotic time when available, whereas the compact sequence component and explicit measurement-density features used the full first 24 hours. For base-model families using eICU development data, meta-learner inputs were generated by five-fold stratified cross-fitting. A 42-input L2-penalized logistic meta-learner combined 16 model probabilities with model-summary, clinical, availability, and measurement-density features.

**Results:** The held-out eICU subset contained 48 positive outcomes (9.39%). The adapted ensemble achieved an area under the receiver operating characteristic curve (AUROC) of 0.841 (95% CI 0.775–0.899) and an average precision (AP) of 0.540 (95% CI 0.399–0.676), with a Brier score of 0.063 and expected calibration error of 0.029. The MIMIC-IV source-only stack achieved an AUROC of 0.782; the paired AUROC difference was 0.059 (95% CI 0.003–0.112). In a separate patient-grouped five-fold out-of-fold sensitivity analysis of 503 linked eICU stays, AUROC was 0.807 (95% CI 0.691–0.910).

**Conclusions:** Target-domain adaptation improved held-out discrimination relative to a source-only stack for sustained antibiotic exposure. Because some model components and measurement-process features used information from the broader first 24 hours, the model should be interpreted as a first-24-hour treatment-association model rather than a strictly prospective pre-treatment infection predictor.

## 1 Background

Spontaneous intracerebral hemorrhage (ICH) is among the most severe forms of acute stroke and remains associated with substantial mortality, long-term disability, and healthcare utilization. Although advances in acute neurological and critical care have improved supportive management, treatment options capable of directly reversing established brain injury remain limited. Early care therefore relies heavily on intensive physiological monitoring, prevention of secondary injury, and timely recognition of complications that arise during hospitalization [1, 2]. Patients requiring intensive care unit (ICU) admission often have impaired consciousness, respiratory compromise, hemodynamic instability, and multiple comorbid conditions, creating a particularly complex clinical environment in which management decisions evolve rapidly during the first days of critical illness.

Infectious complications represent an important component of this early clinical course. Pneumonia, urinary tract infection, bloodstream infection, and other nosocomial infections have been repeatedly described after ICH and are associated with prolonged hospitalization and poorer clinical outcomes [3–5]. In a cohort of patients with spontaneous ICH, nosocomial infection occurred in approximately one quarter of patients, with pneumonia being the most frequent infection [6]. Several features common in severe ICH, including impaired consciousness, mechanical ventilation, invasive monitoring, and systemic inflammatory responses, may both increase infection risk and complicate its recognition. Consequently, antibiotics are often initiated empirically while diagnostic information is still evolving.

Antibiotic exposure in this setting should not, however, be considered equivalent to confirmed infection. Treatment may reflect suspected infection, prophylactic or empiric practice, physiological deterioration, diagnostic uncertainty, and local prescribing patterns. Sustained antibiotic exposure therefore represents a pragmatic treatment-process outcome rather than a microbiologically adjudicated disease endpoint. This distinction is particularly relevant in retrospective electronic health record (EHR) research, where reliable indication-level antibiotic adjudication is often unavailable. Nevertheless, treatment exposure captures an important component of the early ICU trajectory and may be characterized from routinely collected clinical information. Previous studies have demonstrated that longitudinal EHR data can be used to estimate bacterial infection risk and antibiotic-treatment needs in critically ill patients, while more recent work has applied machine learning (ML) to assess antibiotic appropriateness using complex ICU data [7, 8].

Machine-learning approaches have also increasingly been applied to patients with cerebral hemorrhage and ICH. Recent studies using large critical-care databases have evaluated prediction of sepsis and in-hospital mortality using combinations of demographic characteristics, physiological measurements, laboratory variables, and other routinely available clinical features [9–11]. These studies illustrate the potential of EHR-based ML for risk characterization in neurocritical care, but their primary endpoints have generally focused on adverse clinical outcomes such as sepsis or mortality. Treatment-related trajectories have received less attention, and the extent to which models developed in one ICU database remain informative in another healthcare environment is uncertain.

This uncertainty is important because clinical prediction models are sensitive to dataset shift. Differences in case mix, outcome prevalence, measurement frequency, documentation practices, treatment patterns, and variable availability can alter model performance when moving from development to a new setting [12, 13]. ICU-specific evidence similarly shows that externally evaluated models often lose discrimination in new settings and that training across multiple institutions can improve, but does not guarantee, transferability [14, 15]. Generalizability therefore cannot be inferred from within-dataset performance alone, and explicit evaluation of model behavior under changing data environments is necessary [16]. The Medical Information Mart for Intensive Care IV (MIMIC-IV) and eICU Collaborative Research Database provide a useful setting for investigating this problem. MIMIC-IV represents detailed longitudinal data from a single tertiary academic center, whereas eICU contains multicenter criticalcare data from hospitals across the United States [17, 18]. Their differences in patient composition, documentation density, and clinical practice create both an opportunity and a challenge for cross-database modeling.

Rather than treating eICU solely as an untouched external validation dataset, the present study examined whether target-domain information could be incorporated to improve predictive performance in the eICU environment. We developed a library of complementary source and source-plus-target models using MIMIC-IV and an eICU development subset, combined these models through stacked ensembling, and evaluated the resulting pipeline on a held-out eICU subset. The study focused on sustained antibiotic exposure among critically ill patients with ICH, defined pragmatically as antibiotic administration on at least two ICU days. We additionally examined calibration, the contribution of target-domain adaptation and measurement-related features, and robustness to patient-level grouping. Because components of the modeling pipeline incorporated information from the first 24 hours of ICU care and antibiotic exposure could begin within that interval, the task was interpreted as first-24-hour treatment-exposure modeling rather than as strictly prospective pre-treatment prediction of infection.

## 2 Methods

### 2.1 Study design and data sources

This retrospective cross-database modeling study used derived intracerebral hemorrhage cohorts from two publicly available critical-care databases: the Medical Information Mart for Intensive Care IV (MIMIC-IV) and the eICU Collaborative Research Database. MIMIC-IV contains de-identified electronic health record data from patients treated at Beth Israel Deaconess Medical Center, whereas eICU contains multicenter critical-care data collected from hospitals across the United States [17, 18]. Both databases are distributed for research use through established data-access and governance procedures [19].

The analytic design consisted of source-model development using MIMIC-IV, target-domain adaptation using a designated eICU development subset, and final assessment of the fixed adapted pipeline in a held-out eICU evaluation subset. Because eICU data were incorporated during model adaptation and ensemble construction, the primary analysis represented held-out target-domain evaluation after adaptation rather than conventional external validation.

### 2.2 Study population

The analysis began from supplied derived MIMIC-IV and eICU cohort files containing ICU stays associated with intracerebral hemorrhage. Because the original cohort-generation scripts were not available for the present analysis, cohort identification was performed using the supplied derived ICH cohorts rather than reconstructing diagnosis-code extraction procedures from the source databases.

Each observation represented an ICU stay with the information required for the analytic pipeline. MIMIC-IV served as the source cohort for model development. The eICU cohort was stratified by outcome and randomly partitioned at the ICU-stay level into a development subset used for target-domain adaptation and ensemble construction and a held-out evaluation subset reserved for primary performance assessment. The primary partition was not grouped by patient identifier.

Because the derived analytic eICU cohort did not itself contain the longitudinal patient identifier required to identify repeated stays, a separate supplied linkage was used for a patient-grouped sensitivity analysis. Longitudinal patient identifiers could be linked to 503 of the 1,277 eICU stays, representing 452 unique patients. Within this linked subset, five-fold group-aware out-of-fold analysis was performed with patient identity as the grouping variable so that stays from the same patient were assigned to the same fold. The overall study design and analysis workflow are summarized in Figure 1.

**Fig. 1.**
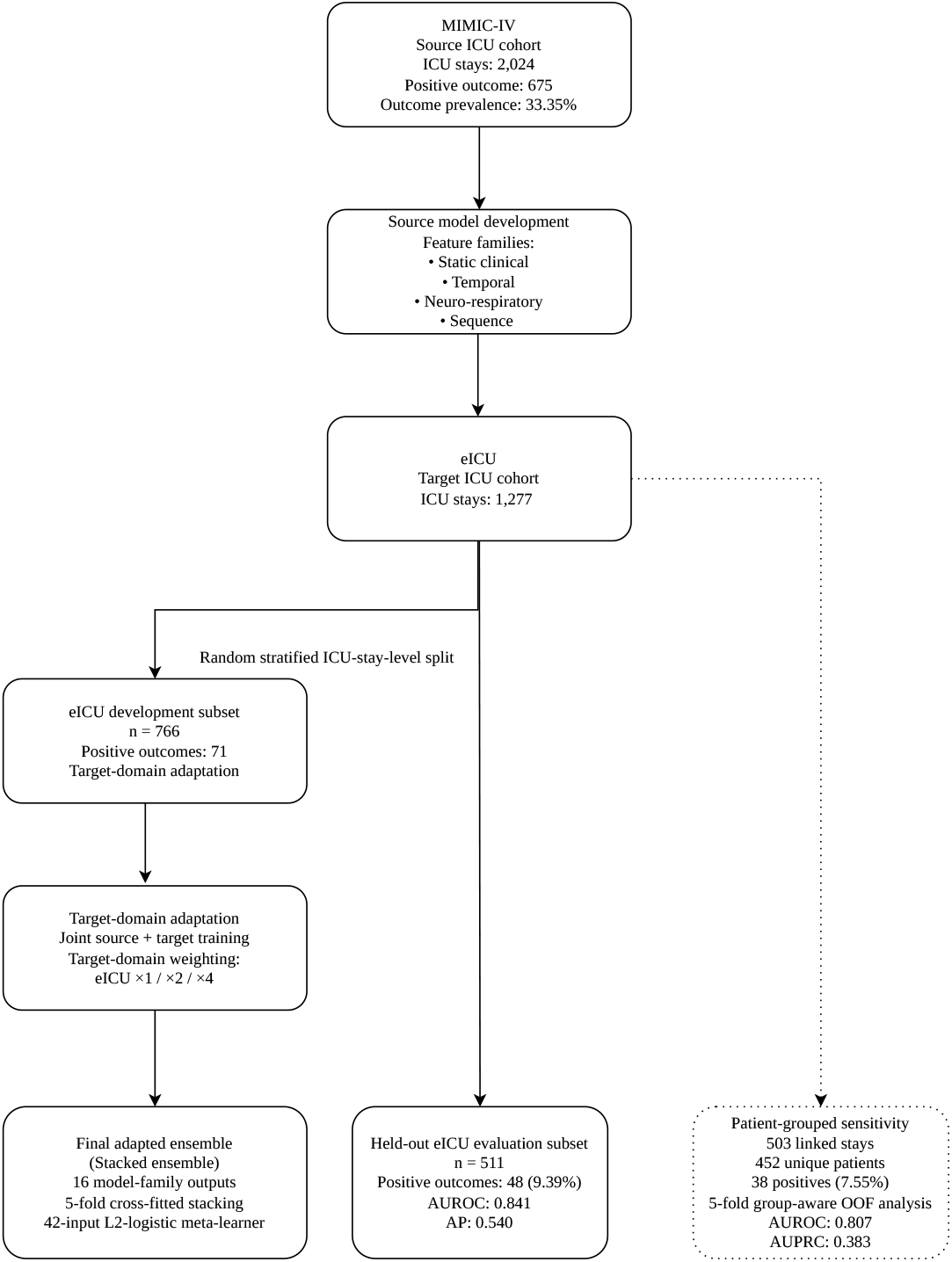
Study design and cross-database adaptation framework. MIMIC-IV served as the source cohort for model development. The eICU target cohort was randomly split at the ICU-stay level into a development subset used for target-domain adaptation and stacked-ensemble construction and a heldout evaluation subset used for final model assessment. For model families using eICU development data, predictions used to train the meta-learner were generated by five-fold stratified cross-fitting. A separate patient-grouped sensitivity analysis was performed among 503 eICU stays that could be linked to longitudinal patient identifiers, representing 452 unique patients.

### 2.3 Outcome definition

The primary outcome was sustained antibiotic exposure during ICU hospitalization. In the eICU cohort, the outcome was defined as antibiotic exposure on at least two ICU-relative days (`abx_days`≥ 2). Antibiotics administered during the first 24 hours after ICU admission could contribute to the outcome if exposure continued into a second ICU day.

In MIMIC-IV, model supervision used the binary source label supplied with the derived cohort. Because the MIMIC-IV and eICU labels were not generated from an identical field, we performed a label-concordance check in MIMIC-IV by reconstructing an analogous exposure proxy based on the number of distinct ICU-relative calendar days containing recorded antibiotic start times and comparing this proxy with the supplied source label.

The available data did not distinguish prophylactic, empiric, and therapeutic antibiotic indications and did not provide adjudicated evidence of infection corresponding to the treatment-exposure outcome. The outcome was therefore interpreted as a treatment-exposure proxy rather than microbiologically confirmed or clinically adjudicated infection.

### 2.4 Feature extraction

Candidate predictors were derived from clinical information recorded during the first 24 hours after ICU admission, with feature-specific handling of antibiotic timing. Feature families were designed to characterize baseline patient status, early physiological evolution, neurologic and respiratory support, observation patterns, and complementary model-level signals.

Baseline variables included age, sex, and comorbidity indicators for hypertension, diabetes mellitus, chronic kidney disease, congestive heart failure, coronary artery disease, atrial fibrillation, chronic obstructive pulmonary disease, liver disease, and malignancy. Physiological and laboratory domains included heart rate, blood pressure, respiratory rate, oxygen saturation, electrolytes, glucose, renal markers, hemoglobin, platelet count, white blood cell count, lactate, and bilirubin. Neurologic and respiratory features included Glasgow Coma Scale components, fraction of inspired oxygen, positive end-expiratory pressure, tidal volume, and ventilator rate.

Repeated clinical measurements were summarized using temporal descriptors including the first and last observed values, minimum, maximum, mean, standard deviation, measurement count, absolute change, and temporal slope. These representations were used to characterize both the level and evolution of clinical measurements during the early ICU course.

Because observation patterns themselves may contain information about clinical status and care intensity, additional care-process features were derived [20, 21]. These included total measurement count, laboratory and vital-sign measurement counts, the proportion of measurements attributable to laboratory testing, the proportion occurring in the later portion of the observation window, and indicators describing availability of individual clinical domains.

Antibiotic timing was handled differently across model components. For the static and temporal tabular model families, timestamped physiological and laboratory observations occurring at or after the recorded first-antibiotic time were excluded from feature construction. Stays without a recorded first-antibiotic time retained the full first-24-hour tabular observation window. In contrast, the compact sequence component and the five explicit measurement-density inputs were calculated from the full first 24 hours after ICU admission. Availability indicators and features derived from a shortened censored window could therefore retain information about observation opportunity and care processes occurring around antibiotic exposure. Availability indicators and features derived from a shortened censored window could therefore retain information about observation opportunity and care processes occurring around antibiotic exposure. The final ensemble was consequently interpreted as a first-24-hour treatment-association model rather than a strictly prospective pre-treatment model.

A summary of the principal predictor and ensemble feature groups is provided in Supplementary Table S2 (Additional file 1).

### 2.5 Machine learning model development and target-domain adaptation

The candidate-model library generated 16 model-family probability outputs representing complementary views of the early ICU course. These included static and temporal CatBoost models [22], neuro-respiratory augmented models, transportable and shift-pruned temporal models, a compact gated recurrent unit (GRU) sequence model, and source-plus-target domain-adapted variants. The model library was designed to combine representations with different levels of temporal detail, feature availability, and sensitivity to cross-database distributional differences.

Target-domain adaptation was performed using the 766-stay eICU development subset while preserving the 511-stay eICU evaluation subset for final assessment. Adapted model families were fitted jointly using MIMIC-IV source data and eICU development data. To vary the influence of target-domain observations during joint training, the eICU development subset was represented at relative weights of one, two, or four times the source-target training contribution.

The final ensemble used stacked generalization [23] with an L2-penalized logistic regression meta-learner. The meta-learner included 42 second-stage inputs: 16 model-level probabilities, 10 summary features derived from those probabilities, 11 prespecified clinical or availability covariates, and five measurement-density features. Probability-summary features included the mean, minimum, maximum, range, and measures of disagreement across candidate-model predictions.

To prevent in-sample base-model predictions from being used to train the metalearner, development-set predictions were generated using five-fold stratified cross-fitting for every base-model family that used eICU development data for adaptation or feature selection. Within each fold, predictions for held-out development stays were obtained from a base model that had not been fitted on those stays. The resulting cross-fitted predictions were then used to train the 42-input meta-learner. After metalearner construction, the corresponding base models were refitted using all available development data and the fixed pipeline was evaluated once on the untouched 511stay eICU evaluation subset. The corrected cross-fitting procedure did not introduce an additional round of model selection.

For comparison with target-domain adaptation, a source-only stacked model was trained entirely using MIMIC-IV data. Both its four constituent base models and its logistic meta-learner were fitted using MIMIC-IV only; the eICU development subset was not used for training, adaptation, feature selection, or meta-learner fitting. The fixed source-only stack was subsequently evaluated on the same held-out eICU evaluation subset as the adapted ensemble.

### 2.6 Model evaluation

Primary model performance was assessed once in the held-out eICU evaluation subset of 511 ICU stays. Discrimination was quantified using the area under the receiver operating characteristic curve (AUROC). Because sustained antibiotic exposure was relatively uncommon in eICU, precision-recall performance was additionally summarized using average precision (AP). Probabilistic performance was evaluated using the Brier score and expected calibration error (ECE). Calibration was also examined graphically by comparing predicted probabilities with observed outcome frequencies.

The final target-domain-adapted ensemble was compared with the fixed MIMIC-IV source-only stack on the same held-out eICU evaluation subset. Performance of individual candidate-model families and the strongest single domain-adapted temporal family was also examined to characterize the incremental contribution of stacked ensembling. Standardized coefficients from the fitted L2-penalized logistic meta-learner were summarized descriptively to illustrate the relative contribution of model-level probabilities, probability-summary features, clinical and availability covariates, and measurement-density features; these coefficients were not interpreted as causal effects or independent feature importance measures.

Robustness to repeated stays from the same patient was evaluated in a separate patient-grouped five-fold out-of-fold analysis among the 503 eICU stays that could be linked to longitudinal patient identifiers. Because this analysis used a different linked subset rather than the primary 511-stay held-out sample, its performance estimates were interpreted as a separate robustness analysis rather than as a paired replacement for the primary evaluation.

### 2.7 Statistical analysis

Cohort sizes, outcome counts, and outcome prevalence were summarized descriptively for the MIMIC-IV source cohort, the eICU development subset, and the held-out eICU evaluation subset. Model performance in the held-out evaluation subset was summarized using AUROC, average precision (AP), Brier score, and expected calibration error (ECE).

Uncertainty in discrimination and precision-recall performance was quantified using nonparametric bootstrap resampling of the held-out eICU evaluation subset. A total of 2,000 bootstrap samples were drawn with replacement, and percentile-based 95% confidence intervals were calculated for the primary AUROC and AP estimates.

Because the final adapted ensemble, the MIMIC-IV source-only stack, and the strongest single domain-adapted temporal family were evaluated on the same held-out eICU stays, differences in AUROC were assessed using paired nonparametric bootstrap resampling. For each bootstrap replicate, the same sampled stay indices were applied to predictions from both models, preserving within-stay pairing. Paired AUROC differences and corresponding 95% bootstrap confidence intervals were calculated for the adapted ensemble versus the source-only stack and for the adapted ensemble versus the strongest single adapted temporal family.

Bootstrap analyses treated the fitted modeling pipelines as fixed and resampled only the held-out evaluation observations; model development, feature selection, target-domain adaptation, cross-fitting, and meta-learner fitting were not repeated within each bootstrap replicate. Accordingly, the resulting intervals quantify uncertainty associated with the held-out evaluation sample rather than uncertainty arising from the full model-development process.

The patient-grouped sensitivity analysis was evaluated separately because it used the 503-stay linked eICU subset rather than the primary 511-stay held-out evaluation sample. Its performance estimates were therefore not used in paired comparisons with the primary analysis.

### 2.8 Use of generative artificial intelligence

A large language model was used to assist with language editing and manuscript restructuring. All methodological descriptions, numerical results, references, and scientific interpretations were independently reviewed and verified by the authors, who take full responsibility for the final manuscript.

## 3 Results

### 3.1 Study cohorts and outcome prevalence

The MIMIC-IV source cohort included 2,024 ICU stays, of which 675 met the supplied positive outcome label, corresponding to a prevalence of 33.35%. The eICU target cohort included 1,277 ICU stays, with 119 positive outcomes (9.32%). Of these, 766 stays were assigned to the development subset used for target-domain adaptation, including 71 positive outcomes (9.27%), and 511 stays were reserved for held-out evaluation, including 48 positive outcomes (9.39%) (Table 1).

**Table 1.** Cohort composition and outcome prevalence in the source and target datasets.

| Cohort | ICU stays | Positive outcomes | Prevalence |
| --- | --- | --- | --- |
| MIMIC-IV source cohort | 2,024 | 675 | 33.35% |
| eICU target cohort | 1,277 | 119 | 9.32% |
| eICU development subset | 766 | 71 | 9.27% |
| eICU held-out evaluation subset | 511 | 48 | 9.39% |

As a label-concordance assessment in MIMIC-IV, the supplied binary source label was compared with a reconstructed proxy based on antibiotic exposure on at least two ICU-relative calendar days. The two definitions disagreed in 3 of 2,024 stays (0.15%), indicating close within-database agreement between the supplied label and the reconstructed proxy. This concordance check did not establish equivalence of outcome construction between MIMIC-IV and eICU, where the supplied `abx days` variable was used directly.

Under the implemented first-antibiotic censoring rule for tabular feature families, 535 of 2,024 MIMIC-IV stays (26.4%) and 92 of 1,277 eICU stays (7.2%) had observation windows truncated before 24 hours. Censoring was more frequent among positive than negative stays in both databases: 51.6% versus 13.9% in MIMIC-IV and 42.9% versus 3.5% in eICU. The MIMIC-IV total included 86 stays with a recorded first-antibiotic time before ICU admission, for which no post-admission tabular measurements were retained under the implemented censoring rule. Stays without a recorded first-antibiotic time retained the full 24-hour tabular observation window.

### 3.2 Primary performance in the held-out eICU evaluation subset

The final target-domain-adapted ensemble achieved an AUROC of 0.841 (95% CI, 0.775–0.899) in the held-out eICU evaluation subset. The corresponding average precision (AP) was 0.540 (95% CI, 0.399–0.676), substantially exceeding the positive-outcome prevalence of 9.39% in the evaluation sample.

Probabilistic performance was characterized by a Brier score of 0.063 and an expected calibration error of 0.029 (Table 2). The calibration curve based on predictions from the final corrected ensemble is shown in Figure 2.

**Table 2.** Primary performance of the final adapted ensemble in the held-out eICU evaluation subset.

| Metric | Estimate |
| --- | --- |
| AUROC | 0.841 (95% CI, 0.775–0.899) |
| Average precision | 0.540 (95% CI, 0.399–0.676) |
| Brier score | 0.063 |
| Expected calibration error | 0.029 |

**Fig. 2.**
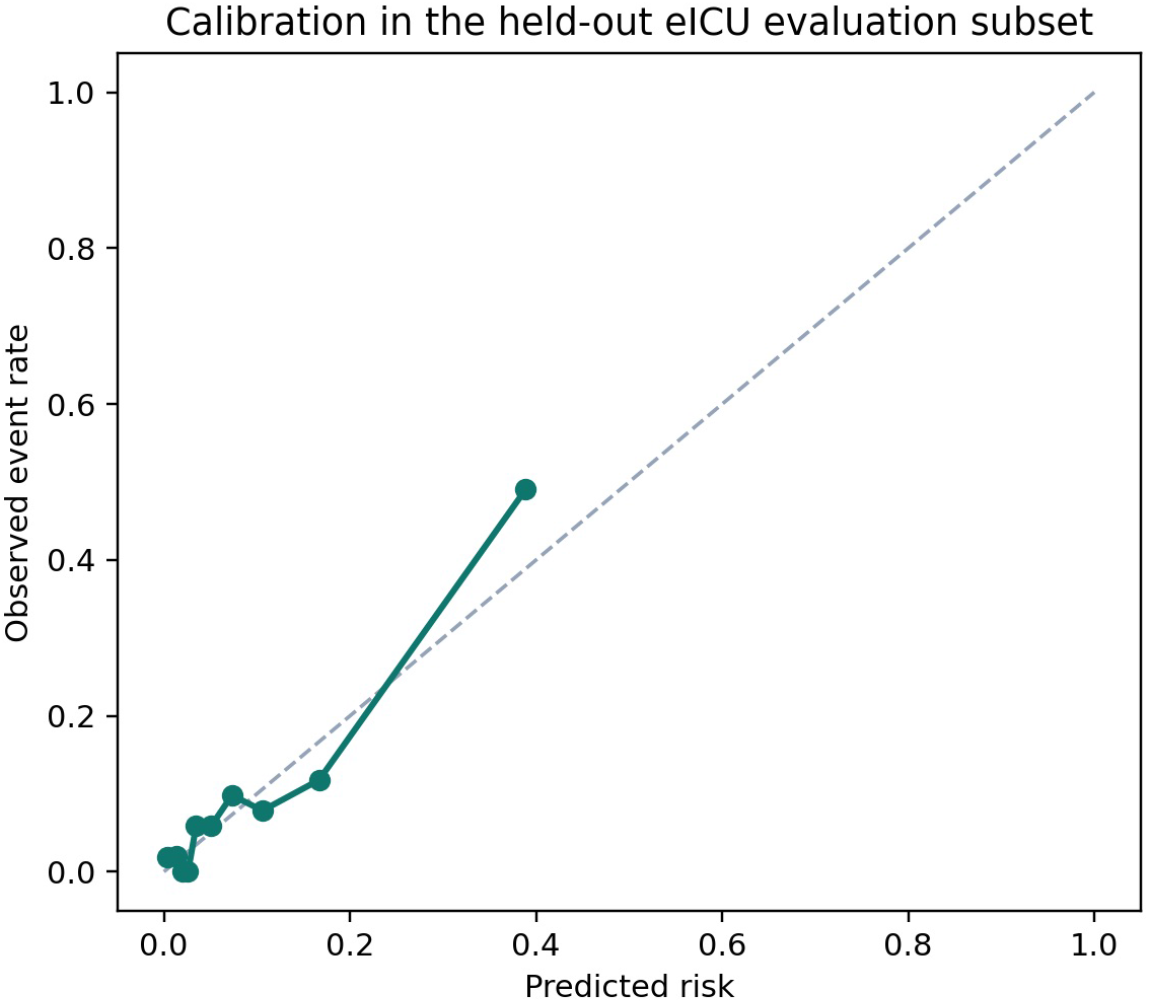
Calibration of the final target-domain-adapted ensemble in the held-out eICU evaluation subset. Predicted probabilities were compared with observed frequencies of sustained antibiotic exposure.

### 3.3 Comparison with source-only and component models

The MIMIC-IV source-only stack achieved an AUROC of 0.782 in the same 511-stay held-out eICU evaluation subset, compared with 0.841 for the final target-domain-adapted ensemble. The paired bootstrap AUROC difference was 0.059 (95% CI, 0.003–0.112). The source-only stack also had a higher Brier score (0.104 vs. 0.063) and expected calibration error (0.172 vs. 0.029) than the adapted ensemble.

Within the broader candidate-model library, the strongest single domain-adapted temporal family achieved an AUROC of 0.805. The paired AUROC difference between the final ensemble and this model was 0.036 (95% CI, −0.017 to 0.085). Thus, the adapted ensemble showed a clear gain over the source-only stack, whereas its incremental discrimination over the strongest single adapted temporal family remained modest and uncertain. Detailed results for benchmark, source-only, and adapted component models are provided in Supplementary Table S1 (Additional file 1).

### 3.4 mPatient-grouped sensitivity analysis

A separate patient-grouped analysis was conducted using the 503 eICU stays that could be linked to longitudinal patient identifiers. These stays represented 452 unique patients and included 38 positive outcomes, corresponding to a prevalence of 7.55%. Five-fold group-aware out-of-fold analysis yielded an AUROC of 0.807 (95% CI, 0.691–0.910) and an AUPRC of 0.383 (95% CI, 0.164–0.613).

This analysis used a linked subset distinct from the primary 511-stay held-out evaluation sample and therefore was not interpreted as a paired replacement for the primary estimate. Nevertheless, discrimination remained above 0.80 after enforcing patient-level grouping.

### 3.5 Contribution of ensemble components

The fitted meta-learner combined model-level predictions with clinical and care-process variables. Among the largest-magnitude standardized coefficients displayed in the final ensemble, the strongest positive coefficient was associated with the logarithm of 24-hour laboratory measurement count. Other positive coefficients included a shift-pruned domain-adapted temporal model, the compact sequence GRU and related sequence-model probability, an indicator for age 60 years or older, female sex, chronic kidney disease, and congestive heart failure. Continuous age and the logarithm of 24-hour vital-sign measurement count had negative coefficients among the leading displayed terms.

These coefficients describe the fitted penalized meta-learner and should not be interpreted as causal effects or as independent measures of feature importance. Several second-stage inputs were correlated, overlapping, or derived from related clinical information. The leading standardized coefficients are shown in Figure 3.

**Fig. 3.**
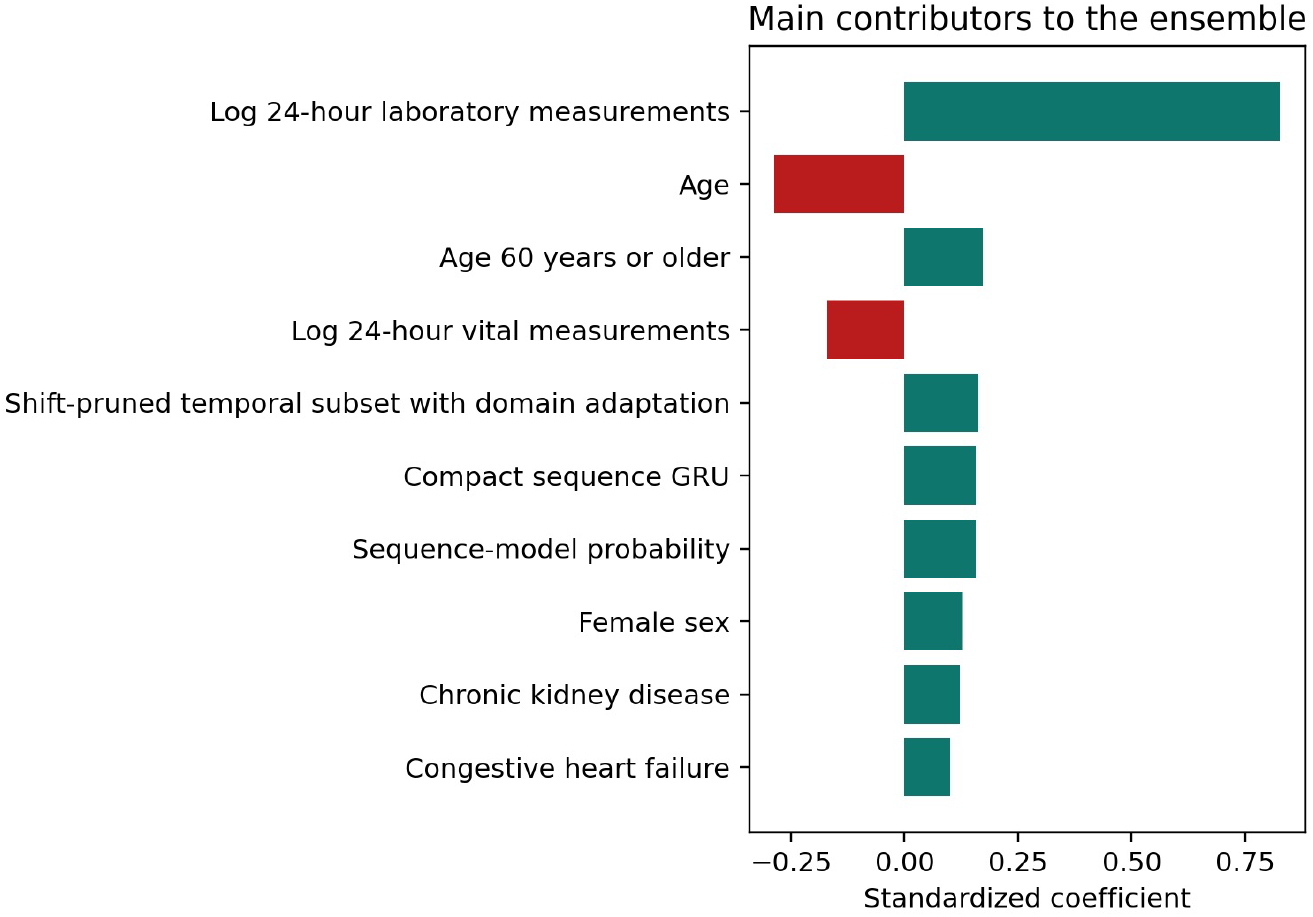
Leading standardized coefficients in the final stacked ensemble. Coefficients are shown for the 42-input L2-penalized logistic meta-learner and should be interpreted descriptively because several model-level, clinical, and care-process inputs are correlated.

## 4 Discussion

In this cross-database study of critically ill patients with intracerebral hemorrhage, the final target-domain-adapted ensemble achieved an AUROC of 0.841 and an average precision of 0.540 in the held-out eICU evaluation subset, with a Brier score of 0.063 and an expected calibration error of 0.029. The fixed MIMIC-IV source-only stack achieved an AUROC of 0.782 in the same evaluation cohort, and the paired AUROC difference was 0.059 (95% CI, 0.003–0.112). A separate patient-grouped analysis yielded an AUROC of 0.807. Together, these findings suggest that incorporating target-domain data improved held-out discrimination relative to a source-only model while preserving useful predictive signal under patient-level grouping.

The clinical meaning of this task requires careful interpretation. Infectious complications are common after ICH and are associated with additional morbidity, prolonged hospitalization, and adverse outcomes [3–6]. Patients with severe neurological injury may be particularly susceptible to pneumonia and other nosocomial infections because of impaired consciousness, aspiration risk, mechanical ventilation, invasive procedures, and prolonged ICU exposure. At the same time, systemic inflammatory responses after acute brain injury can complicate the distinction between infectious and noninfectious deterioration, and antibiotics may be initiated empirically before diagnostic certainty is available. The outcome in the present study should therefore be interpreted as a treatment-exposure phenotype reflecting both patient condition and clinical management rather than as microbiologically confirmed or clinically adjudicated infection.

Previous machine-learning studies in ICH have largely focused on more conventional adverse outcomes, including sepsis and mortality [9–11]. Related work in general ICU populations has shown that longitudinal electronic health record data can be used to estimate bacterial infection risk, antibiotic-treatment requirements, or antimicrobial-treatment appropriateness [7, 8]. The present study differs in both outcome and design. First, sustained antibiotic exposure was modeled rather than an adjudicated infectious diagnosis or treatment-appropriateness label. Second, the main methodological question concerned cross-database adaptation: whether a modeling pipeline developed primarily from one ICU database could be adapted using limited target-domain data and subsequently evaluated in a held-out target-domain sample. This distinction is important because predictive models can degrade substantially under dataset shift [12, 13]. Positive-outcome prevalence was 33.35% in MIMIC-IV but approximately 9.3% in eICU. Part of this difference may reflect label-definition drift: MIMIC-IV used a supplied binary source label, whereas eICU used the supplied `abx days` variable with sustained exposure defined as antibiotic administration on at least two ICU days. Although a reconstructed MIMIC-IV proxy disagreed with the supplied source label in only 3 of 2,024 stays, this within-database concordance does not establish equivalence of outcome construction between MIMIC-IV and eICU. Differences in case mix, treatment practice, medication documentation, ICU organization, and cohort construction may also contribute to the prevalence gap. These sources of heterogeneity reinforce the broader observation that strong within-dataset performance alone is insufficient evidence of transportability [14–16]. Related cross-database ICU studies using MIMIC and eICU have likewise emphasized external evaluation and calibration assessment across heterogeneous data environments [24, 25].

The higher performance of the adapted ensemble relative to the MIMIC-IV source-only stack is therefore methodologically relevant. The source-only comparator was trained entirely using MIMIC-IV, whereas the adapted pipeline incorporated the 766-stay eICU development subset through joint source-plus-target modeling and target-domain weighting. In the same 511-stay held-out eICU evaluation subset, the adapted ensemble achieved an AUROC 0.059 higher than the source-only stack, with a paired bootstrap confidence interval that did not include zero. The adapted ensemble also showed lower Brier score and expected calibration error. These results support the value of incorporating target-domain information in this setting, but they should not be interpreted as evidence that the specific adaptation strategy used here is universally optimal. Because eICU development data contributed to model adaptation and ensemble construction, the final evaluation represents held-out target-domain assessment after adaptation rather than conventional external validation.

The incremental value of stacking beyond the strongest single adapted model was more modest. The best single domain-adapted temporal family achieved an AUROC of 0.805, corresponding to a paired difference of 0.036 relative to the final ensemble (95% CI, −0.017 to 0.085). The confidence interval included zero, indicating uncertainty regarding whether the stacked architecture consistently improves discrimination beyond the strongest individual adapted family. The contribution of the ensemble may therefore lie less in model complexity itself than in combining complementary representations of the early ICU course. This interpretation is also consistent with the fitted meta-learner, which combined model-level probabilities with low-dimensional summary, clinical, availability, and measurement-process features rather than relying on a single dominant predictive representation.

Care-process information remains an important component of the final model. Among the largest standardized meta-learner coefficients, 24-hour laboratory measurement intensity contributed prominently, alongside several model-level and clinical terms. Such observation patterns may encode both patient severity and healthcare processes [20, 21]. Importantly, feature construction was not uniformly restricted to information preceding antibiotic exposure. For static and temporal tabular model families, timestamped observations at or after the first recorded antibiotic time were excluded when such timing was available. In contrast, the compact sequence component and five explicit measurement-density inputs were calculated over the full first 24 hours. In addition, shorter censoring windows and availability patterns could indirectly encode differences in observation opportunity. First-antibiotic censoring occurred disproportionately among positive stays in both databases. Accordingly, the final pipeline is better interpreted as a first-24-hour treatment-association model than as a strictly prospective pre-treatment prediction model. Model coefficients involving measurement intensity or availability should therefore not be interpreted as purely biological predictors.

The patient-grouped sensitivity analysis provides additional context for the primary stay-level evaluation. Longitudinal patient linkage was available for 503 eICU stays representing 452 unique patients and 38 positive outcomes. Five-fold group-aware out-of-fold analysis produced an AUROC of 0.807 and an AUPRC of 0.383. Because this linked subset differed from the primary 511-stay held-out evaluation cohort, these estimates cannot be interpreted as a paired correction of the primary result. Nevertheless, preservation of an AUROC above 0.80 after enforcing patient-level grouping provides some reassurance that the main discrimination signal was not solely explained by repeated stays from the same patient. The wider uncertainty in this analysis also reflects the smaller number of positive outcomes and the restricted linkage coverage.

The potential clinical role of the model should remain conservative. Prediction of sustained antibiotic exposure does not determine whether antibiotic treatment was necessary, appropriate, or beneficial and should not be interpreted as a recommendation to initiate or continue antimicrobial therapy. Instead, the present model may be useful for characterizing treatment trajectories and for examining how early physiology, neurological and respiratory status, model-derived risk signals, and care processes relate to antibiotic exposure across heterogeneous ICU environments. Evaluation for antimicrobial stewardship or bedside decision support would require outcomes incorporating microbiological evidence, clinical infection adjudication, antibiotic indication, and prospectively defined treatment decisions.

Several limitations should be acknowledged. First, the outcome was an antibiotic-exposure proxy and did not distinguish prophylactic, empiric, and therapeutic treatment or confirm infection. Differences in outcome construction between MIMIC-IV and eICU remain a potential source of label-definition drift despite the close internal concordance observed within MIMIC-IV. Second, the original cohort-generation scripts used to derive the supplied ICH cohorts were unavailable, preventing independent reconstruction of the initial diagnosis-code-based cohort-identification step. Third, the feature-generation process created unequal observation opportunities: tabular features could be censored at first antibiotic administration, whereas the sequence component and measurement-density inputs used the full first 24 hours. Because censoring occurred more frequently among positive stays, some predictive information may reflect treatment timing or contemporaneous care rather than purely antecedent physiology. Fourth, the primary eICU development and evaluation split was performed at the ICU-stay rather than patient level, and longitudinal linkage was available for only 503 stays; the patient-grouped sensitivity analysis therefore used a smaller and different subset. Fifth, because eICU development data were used for target-domain adaptation, the study does not constitute fully independent external validation.

Additional limitations relate to sample size and model complexity. The final 42input L2-penalized logistic meta-learner was fitted in the 766-stay eICU development subset containing 71 positive outcomes. Although most second-stage inputs were model probabilities, low-dimensional summaries, or prespecified clinical and availability covariates rather than a new high-dimensional raw feature set, the limited event count relative to the number of meta-learner inputs may still contribute to coefficient instability and optimism. Only 48 positive outcomes were available in the primary held-out evaluation subset, and 38 were available in the patient-linked sensitivity subset, resulting in relatively wide confidence intervals, particularly for precision-recall performance. The candidate-model and adaptation search was also not repeated within a fully nested validation framework; bootstrap intervals therefore quantify uncertainty in the fixed final pipeline rather than uncertainty arising from the complete development and model-selection process. Finally, harmonized APACHE or complete SOFA scores were not available from the supplied eICU export at the matched evaluation point, precluding a same-cohort comparison with established bedside severity scores.

Despite these limitations, the study illustrates that target-domain adaptation can retain and improve useful predictive information when a modeling pipeline is transferred between heterogeneous ICU databases. It also demonstrates why cross-database evaluation should consider not only discrimination but also calibration, patient grouping, label construction, and the contribution of healthcare-process information. For treatment-related outcomes in particular, separating antecedent biological risk from concurrent clinical management will be important in determining whether similar models can support future prospective clinical investigation.

## 5 Conclusions

In critically ill patients with intracerebral hemorrhage, a target-domain-adapted ensemble showed higher held-out discrimination than a fixed MIMIC-IV source-only stack for sustained antibiotic exposure in eICU. The adapted ensemble achieved an AUROC of 0.841, and the paired AUROC difference relative to the source-only comparator was 0.059 (95% CI, 0.003–0.112). Performance remained above 0.80 in a separate patient-grouped sensitivity analysis.

These findings support the value of target-domain adaptation when transferring ICU prediction models across heterogeneous databases. However, because the outcome reflected treatment exposure rather than adjudicated infection and some model components incorporated information from the broader first 24 hours of ICU care, the final pipeline should be interpreted as a treatment-association model rather than as a strictly prospective pre-treatment infection predictor or a direct antimicrobial decision tool.

## Data Availability

The MIMIC-IV and eICU Collaborative Research Database datasets analyzed in this study are available to credentialed investigators through PhysioNet (https://physionet.org/content/mimiciv/ and https://physionet.org/content/eicu-crd/) under their respective data-use requirements. The present analysis was performed using derived intracerebral-hemorrhage cohort files generated from these source databases. These derived patient-level files are not redistributed because they originate from credentialed-access clinical databases.

## Abbreviations

AP: average precision
APACHE: Acute Physiology and Chronic Health Evaluation
AUPRC: area under the precision-recall curve
AUROC: area under the receiver operating characteristic curve
ECE: expected calibration error
EHR: electronic health record
GRU: gated recurrent unit
ICH: intracerebral hemorrhage
ICU: intensive care unit
ML: machine learning
MIMIC-IV: Medical Information Mart for Intensive Care
IV: PEEP positive end-expiratory pressure
SOFA: Sequential Organ Failure Assessment.

## Supplementary information

Additional file 1. File format: PDF. Title: Supplementary Information. Description: Contains Supplementary Table S1 (performance of representative benchmark, source-only, and adapted models) and Supplementary Table S2 (principal predictor and ensemble feature groups).

## Declarations

### Ethics approval and consent to participate

This study was a secondary analysis of de-identified data from the MIMIC-IV and eICU Collaborative Research Database and involved no direct patient contact or intervention. The collection of patient information and creation of the MIMIC-IV research resource were reviewed by the Institutional Review Board of Beth Israel Deaconess Medical Center, which approved the data-sharing initiative and granted a waiver of informed consent. The eICU Collaborative Research Database contains de-identified retrospective critical-care data and is exempt from institutional review board approval for secondary research use because of the retrospective design, absence of direct patient intervention, and privacy safeguards meeting Health Insurance Portability and Accountability Act Safe Harbor standards. No additional individual informed consent was required for the present secondary analysis.

### Consent for publication

Not applicable. This study used de-identified secondary data and contains no individually identifiable patient information.

### Availability of data and materials

The MIMIC-IV and eICU Collaborative Research Database datasets analyzed in this study are available to credentialed investigators through PhysioNet under their respective data-use requirements [17–19]. The present analysis was performed using derived intracerebral-hemorrhage cohort files generated from these source databases. These derived patient-level files are not redistributed with the manuscript because they originate from credentialed-access clinical databases.

### Authors’ contributions

SP performed the primary data analysis, feature engineering, model development, and visualization. YS led methodological review, interpretation of the analyses, manuscript restructuring, and preparation and revision of the manuscript. JS, MH, and NTK contributed to study methodology, interpretation of the results, and manuscript preparation and revision. GP and KA contributed to methodological and clinical review and critically revised the manuscript. MP contributed to study conceptualization and interpretation, supervised the project, and critically revised the manuscript. SP, YS, JS, MH, and NTK contributed equally to this work. All authors read and approved the final manuscript.

## Acknowledgements

Not applicable.

