## Additional file 1: Supplementary Tables S1 and S2 for "Cross-database adaptation of machine learning models for predicting sustained antibiotic exposure among patients with intracerebral hemorrhage in intensive care units"

### Additional file 1: Supplementary Information

#### Supplementary Table S1

Table S1: Performance of representative benchmark, source-only, and adapted models in the held-out eICU evaluation subset.

| Model | AUROC | Brier score | ECE |
|---|---|---|---|
| Admission-factor logistic comparator | 0.730 | 0.078 | – |
| Static clinical summary | 0.737 | 0.121 | 0.203 |
| Temporal physiologic summary | 0.736 | 0.112 | 0.187 |
| Compact sequence GRU | 0.767 | 0.142 | 0.233 |
| Static plus neuro-respiratory model | 0.787 | 0.114 | 0.189 |
| MIMIC-IV source-only stack | 0.782 | 0.104 | 0.172 |
| Best single domain-adapted temporal family | 0.805 | 0.073 | 0.054 |
| Final target-domain-adapted ensemble | **0.841** | **0.063** | **0.029** |

AUROC, area under the receiver operating characteristic curve; ECE, expected calibration error; GRU, gated recurrent unit. The table shows representative benchmark and component models rather than all 16 model-family probability outputs used by the final meta-learner. The paired AUROC difference between the final adapted ensemble and the MIMIC-IV source-only stack was 0.059 (95% CI, 0.003–0.112). The paired AUROC difference between the final ensemble and the strongest single domain-adapted temporal family was 0.036 (95% CI, −0.017 to 0.085).

#### Supplementary Table S2

Table S2: Principal feature groups considered during model development and target-domain adaptation.

| Feature group | Variables or representations |
|---|---|
| Demographic and comorbidity features | Age, sex, hypertension, diabetes mellitus, chronic kidney disease, congestive heart failure, coronary artery disease, atrial fibrillation, chronic obstructive pulmonary disease, liver disease, and malignancy. |
| Physiological and laboratory features | Heart rate, blood pressure, respiratory rate, oxygen saturation, electrolytes, glucose, renal markers, hemoglobin, platelet count, white blood cell count, lactate, and bilirubin. |
| Temporal summaries | First and last observed values, minimum, maximum, mean, standard deviation, measurement count, absolute change, and temporal slope. |
| Neurologic and respiratory features | Glasgow Coma Scale components, fraction of inspired oxygen, positive end-expiratory pressure, tidal volume, and ventilator rate. |
| Observation and measurement-density features | Total measurement count, laboratory and vital-sign measurement counts, laboratory proportion, late-window measurement proportion, and indicators describing availability of individual clinical domains. |
| Model-level ensemble features | Base-model probabilities and summary statistics across model probabilities, including mean, minimum, maximum, range, and measures of disagreement. |

For static and temporal tabular model families, timestamped physiological and laboratory observations occurring at or after the recorded first-antibiotic time were excluded from feature construction. Stays without a recorded first-antibiotic time retained the full first-24-hour tabular observation window. The compact sequence component and five explicit measurement-density inputs were calculated from the full first 24 hours after ICU admission.
